# The potential health impact and healthcare cost savings of different sodium reduction strategies in Canada

**DOI:** 10.1101/2023.11.02.23297997

**Authors:** Nadia Flexner, Amanda C. Jones, Ben Amies-Cull, Linda Cobiac, Eduardo Nilson, Mary R. L’Abbe

## Abstract

**Background:** High dietary sodium is the main dietary risk factor for non-communicable diseases due to its impact on cardiovascular diseases, the leading cause of death globally. The Government of Canada has taken measures to reduce average dietary sodium intakes, such as setting voluntary sodium reduction targets for packaged foods and recently approving regulations mandating ‘high in’ front-of-pack labeling (FOPL) symbols.

**Objectives:** To estimate the number of avoidable ischemic heart disease (IHD) and stroke incidence cases, and their associated healthcare cost and Quality-Adjusted Life Year (QALY) savings resulting from different sodium reduction strategies and recommendations in Canada.

**Methods:** We used the PRIMEtime model, a proportional multi-state lifetable model. Outcomes were modeled over the lifetime of the population alive in 2019, at a 1.5% discount rate, and from the public healthcare system perspective. Nationally representative data were used as inputs for the model.

**Results:** Fully meeting Health Canada’s sodium reduction targets was estimated to prevent 219,490 (95% UI, 73,409–408,630) cases of IHD, and 164,435 (95% UI, 56,121–305,770) strokes. This led to a gain of 276,185 (95% UI, 85,414–552,616) QALYs, and healthcare costs savings of CAD$ 4,212(95% UI, 1,303–8,206) million over the lifetime of the 2019 cohort. Sodium reduction intake through FOPL regulations has the potential to prevent between 35,930 (95% UI, 8,058– 80,528) and 124,744 (95% UI, 40,125–235,643) cases of IHD, and between 26,869 (95% UI, 5,235–61,621) and 93,129 (95% UI, 30,296–176,014) strokes. This results in QALY gains ranging from 45,492 (95% UI, 10,281–106,579) to 157,628 (95% UI, 46,701–320,622), and healthcare costs savings ranging from CAD$ 695 (95% UI, 160–1,580) to CAD$ 2,415 (95% UI, 722–4,746) million over the lifetime of the 2019 Canadian cohort. Greater health and healthcare costs gains were estimated if Canadians were to meet the population-level sodium intake recommendations of the World Health Organization (2,000 mg/day) and the Adequate Intake recommendation (1,500 mg/day). All sodium reduction strategies tested were cost saving.

**Conclusions:** Reducing population-level sodium intakes is feasible and has the potential to improve health outcomes and save healthcare costs in Canada. From interventions tested, most health and healthcare costs gains were attributed to fully meeting sodium reduction targets, which highlights the importance of changing the voluntary nature of these targets to mandatory. A combination of strategies, mandatory sodium reduction targets and implementation of the ‘high in’ FOPL symbol would provide the most benefit from a public health standpoint.

## 1. Introduction

Diets high in sodium are the main dietary risk factor for non-communicable diseases (NCDs)(1) due to their impact on cardiovascular diseases (CVDs)(2, 3), the leading cause of death globally and a major contributor to disability, particularly in relation to diseases such as ischemic heart disease (IHD) and stroke(4). In Canada, CVDs (mostly IHD and strokes) rank as the second leading cause of death (after cancers), accounting for 25% of all deaths each year(5). The World Health Organization (WHO) broadly considers reducing population-level dietary sodium intake to be a cost-effective measure that could save many lives by decreasing and preventing the burden of diet-related NCDs, mainly from CVDs, and save costs in the long run(6, 7).

The WHO recommends consuming less than 2,000 mg/day of sodium (5 g/day of salt)(8); however, intake levels around the world largely exceed recommendations(8). Canada is not an exception, as recent sodium intake estimations showed that Canadians consume an average of 2,760 mg/day of sodium(9, 10), with 63.9% of Canadian adults exceeding the recommendation for Chronic Disease Risk Reduction (CDRR) levels (2,300 mg/day) and only 9.4% of Canadian adults meeting the Adequate Intake (AI) levels (1,500 mg/day)^10^.

Reducing sodium intake is a WHO ‘best buy’ recommendation for the prevention and control of NCDs(6). In its *SHAKE Technical Package for Salt Reduction*(11), WHO recommends ideally implementing a joint combination of measures to reduce population-level sodium intakes. These measures include monitoring salt use (e.g., intakes, salt content in foods), promote food reformulation to contain less salt by setting salt targets levels in foods, adopting interpretive front-of-pack labeling systems, and restricting marketing of foods ‘high in’ salt, among others(11).

The Canadian government has implemented and adopted several evidence-based policies to make the healthier choice easier for all Canadians, as described in the *Canadian Healthy Eating Strategy* (2016)(12). As part of this comprehensive strategy, Health Canada committed to ongoing collaboration with stakeholders in the food industry to reduce sodium levels in packaged foods through updating the voluntary sodium reduction targets for processed foods (2020-2025)(13). It is worth noting that progress evaluations of the previous (2012-2016) set of voluntary sodium reduction targets have only shown modest results(14), and only minor revisions have been made to the latest set of targets (2020-2025)(13). In addition, Health Canada recently approved front-of-pack labeling (FOPL) regulations that will require packaged foods that meet or exceed established thresholds for nutrients-of-concern, including sodium, to display a ‘high in’ FOPL nutrition symbol, to come into effect by January 2026(15).

Estimating the impact of public health polices, such as the aforementioned, could be challenging or simply not feasible using traditional epidemiological research methods such as cohort studies, randomized control trials or natural experiments. Policy scenario modeling has proven to be an appropriate and strategic tool for estimating the health and economic impact of public health policies prior to actual implementation(16). Specifically, NCD policy scenario modeling allows us to predict the cumulative health outcomes of interventions over the long term and across the entire population; in addition to providing evidence on what the magnitude would be to take action or not by comparing different policy scenarios with a ‘business as usual’ scenario(17).

A systematic review of previous policy scenario modeling studies have demonstrated the cost-effectiveness of reducing dietary sodium intake(18). Studies in the Canadian context have estimated the economic and health impact of reducing sodium intake to recommended levels when compared to previous sodium intake levels (i.e., 3,800 mg/day) using data from CCHS-Nutrition 2004(19). More recently, using data from CCHS-Nutrition 2015, we previously estimated the number of CVDs deaths that could be averted or delayed if Canadians were to reduce their sodium intake to recommended levels(10, 19), and as a result of implementing ‘high in’ FOPL regulations(20, 21). However, to the best of our knowledge, no study in Canada has previously estimated the health impact and healthcare cost savings from ‘real world’ policy scenario options to reduce population level dietary sodium intakes (i.e., food reformulation and mandatory ‘high in’ FOPL symbol).

In this study, we aimed to provide policymakers and stakeholders with evidence supporting policy options to reduce excess dietary sodium intakes in Canada. We estimated the number of avoidable ischemic heart disease and stroke incidence cases, and their associated healthcare cost, and Quality-Adjusted Life Year (QALY) savings resulting from 1) meeting sodium intake recommended levels, 2) meeting sodium reduction targets for packaged foods established by Health Canada, and 3) implementing ‘high in’ front-of-pack labeling regulations in Canada.

## 2. Materials and methods

We conducted a cost-utility analysis using a public healthcare system perspective. The potential health and healthcare costs impact of reducing dietary sodium intake were modelled by comparing ten counterfactual scenarios (described below in section *‘Counterfactual scenarios modeled’*) to a ‘business as usual’ or ‘baseline’ scenario. Outcomes were modeled over the lifetime of the Canadian population alive in 2019, and a 1.5% discount rate was applied to both health gains and healthcare costs, as recommended by the Canadian Agency for Drugs and Technologies in Health (CADTH)(22).

### 2.1 PRIMEtime model: a proportional multi-state lifetable model

The PRIMEtime model, an established and robust proportional multi-state lifetable model, was used in this study(23, 24). PRIMEtime is an epidemiological model tool, developed by researchers at the University of Oxford, that has been widely applied to a range of different diet and obesity policy scenarios in the UK and other countries(24-31). This model has been designed for policy scenario evaluation and can be utilized to assess future effects of policies in order to help policymakers set priorities for action. In a world of limited resources, governments are increasingly required to properly assess the cost-effectiveness of interventions to strategically allocate their resources. The PRIMEtime model is a tool for policy support and prioritization that under the same set of assumptions can help compare different policy options by assessing the cost-effectiveness of interventions.

The PRIMEtime model is comprised of the following interconnected modules: a risk factor exposure module, a series of disease models, and a lifetable. Detailed methods of the model have been published elsewhere(24). Briefly, PRIMEtime uses epidemiological data collected mainly from meta-analyses of randomized control trials or prospective cohort studies to parameterize the relationships between risk factors and NCDs. This can be modeled as direct effects of risk factors or via their intermediate risk factors (i.e., blood pressure, BMI, or total cholesterol), where appropriate. The Canadian PRIMEtime Salt model includes the two main CVDs associated with high consumption of salt/sodium, IHD and stroke. These diseases are modeled in PRIMEtime in a three-state Markov model in which the population is either in a disease-free state, a diseased state, or dead. Then, the lifetable combines these simulated changes in population health and healthcare system as a result of changes in disease epidemiology.

We adapted the PRIMEtime model to the Canadian context by collecting, processing, and analyzing Canadian-specific and nationally representative data (i.e., dietary sodium intakes, population demographics, disease epidemiology and healthcare costs). **Table 1** shows the data sources used as inputs for the PRIMEtime model in Canada. A closed cohort analyses was performed, taking the Canadian adult population as a baseline (≥19 y) over the lifetime of the cohort or until they reach 100y of age.

**Table 1.**
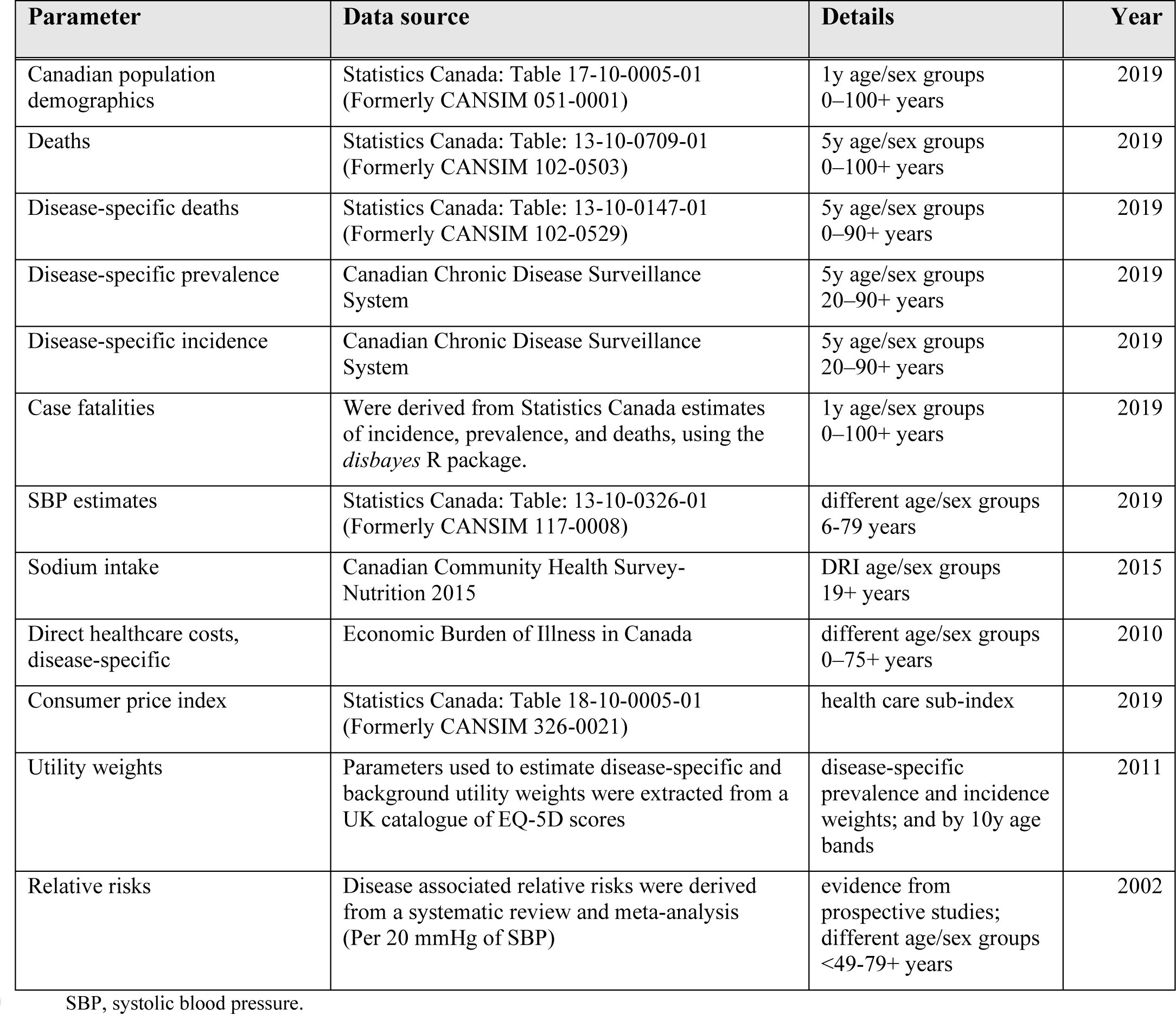
Key data sources used for the application of the PRIMEtime Salt model in Canada.

### 2.2 Model input parameters

#### Population inputs

Canadian population size and all-cause mortality data by 1-year age/sex groups were included in the model. Data was obtained from the publicly available Statistics Canada CANSIM tables for the year 2019 **(Table 1, Supplementary Table S1)**. All-cause mortality data was only available in 5-year age/sex groups; thus, further processing was required. Data was interpolated to 1-year using a temporal disaggregation method(32).

#### Disease epidemiology

Data on incidence, prevalence and disease-specific mortality were collected for ischemic heart disease and stroke for the year 2019 **(Table 1)**. First, we prioritized alignment of WHO International Statistical Classification of Diseases and Related Health Problems 10th Revision (ICD-10) codes(33) between disease epidemiology and healthcare cost data (described below). Disease epidemiology data was only available in 5y age/sex groups up to age 90+ for all parameters; thus, further processing was required. Data was extrapolated to age 100+ using a polynomial trend line (**Supplementary Table S2**). Then, health data estimates were interpolated to 1y age/sex groups using a temporal disaggregation method to obtain smooth disaggregated counts, while maintaining the aggregated total(32).

Case fatality rates were derived from cause-specific incidence, prevalence and mortality data, together with population size using a Bayesian approach, the *disbayes* optimization method built on the Stan software (*disbayes* package available in R)(32, 34). It was assumed that case fatality was constant for all ages below 35. The *disbayes* package estimates case fatality using a three-state transition process of disease free, disease, or death. The *disbayes’* approach and assumptions are consistent with the ones underpinning disease simulation in the PRIMEtime model (**Supplementary Table S1)**.

Mean systolic blood pressure (SBP) estimates for the year 2019 were acquired. Data was available by sex (male, female) and age group (6-11 years, 12-19, 20-39, 40-59, 60-79) **(Table 1)**. As SBP measurements were not available for individuals aged 80 years and older, we assumed the same values as those observed for 60-79 years age/sex groups for individuals in the 80+ years group.

#### Dietary sodium intake

This study used data from the Canadian Community Health Survey (CCHS)-Nutrition 2015 to estimate baseline and counterfactual dietary sodium intakes for Canadian adults (≥19 y). CCHS-Nutrition 2015 is a nationally representative, cross-sectional health and nutrition survey that utilized 24-hour (24h) dietary recalls to collect data on food and beverage intake across the 10 Provinces of Canada (aged 1y or older)(35). We used both available days of 24h dietary recalls to estimate usual dietary sodium intakes for adults by Dietary Reference Intakes (DRI)(36) age/sex groups (i.e., males and females ages 19-30, 31-50, 51-70, 71+), methods that have been previously published by our research group(10, 20, 21).

#### Counterfactual scenarios modeled

#### Meeting sodium intake recommended levels (Scenarios 1 and 2)

We modeled potential health and healthcare costs gains from meeting WHO sodium intake recommendations of 2,000 mg/day and the AI recommendations of 1,500 mg/day. The proportion needed to meet these recommendations from the current sodium intakes (2,758 mg/day) was calculated and applied to each DRI age/sex group to build our counterfactual scenarios^10^ **(Table 2, Supplementary Table S3)**.

**Table 2.** Sodium reduction counterfactual scenarios.

| Counterfactual scenarios |  | Based on | Parameters modeled | Δ Sodium* | Δ Salt* |
| --- | --- | --- | --- | --- | --- |
| **Meeting sodium intake recommendations** | S1: AI sodium intake recommendations (1,500 mg/day) | Flexner N, Christoforou A, et al (2023) (10) | Estimated sodium intakes by DRI age/sex group | - 1,258 mg/day | - 3.14 g/day |
|  | S2: WHO sodium intake recommendations (2,000 mg/day) | Flexner N, Christoforou A, et al (2023) (10) | Estimated sodium intakes by DRI age/sex group | - 758 mg/day | - 1.89 g/day |
|  | S3: Sodium reduction targets for packaged foods, reformulation (2,300 mg/day) | Flexner N, Christoforou A, et al (2023) (10) | Food & beverages with a sodium target | - 459 mg/day | - 1.15 g/day |
| **Implementing ‘high in’ FOPL regulations (food and beverage purchases)** | S4: Changes in food & beverage purchases (Chilean FOPL experience) (20) | Taillie L, et al (2021) (38) | Changes in sodium content of food & beverage purchases | - 128 mg/day | - 0.32 g/day |
|  | S5: Based on WHO criteria to estimate cost-effectiveness of FOPL policies (20) | WHO (2022) (40) | Changes in sodium content of food & beverage purchases | - 174 mg/day | - 0.44 g/day |
|  | S6: Based on a FOPL systematic review and network meta-analysis (20) | Song J, et al (2021) (39) | Changes in sodium content of food & beverage purchases | - 212 mg/day | - 0.53 g/day |
| **Implementing ‘high in’ FOPL regulations (food substitution)** | S7: Based on food substitution for 30% of CCHS-Nutrition 2015 adult participants | Flexner N, et al (2023) (21) | Food substitution with a healthier food alternative | - 73 mg/day | - 0.18 g/day |
|  | S8: Based on food substitution for 50% of CCHS-Nutrition 2015 adult participants | Flexner N, et al (2023) (21) | Food substitution with a healthier food alternative | - 131 mg/day | - 0.33 g/day |
|  | S9: Based on food substitution for 70% of CCHS-Nutrition 2015 adult participants | Flexner N, et al (2023) (21) | Food substitution with a healthier food alternative | - 182 mg/day | - 0.45 g/day |
|  | S10: Based on food substitution for all CCHS-Nutrition 2015 adult participants | Flexner N, et al (2023) (21) | Food substitution with a healthier food alternative | - 259 mg/day | - 0.65 g/day |

\*These values are overall intervention effect. Intervention effect by DRI age/sex group, as imputed in the PRIMETIME model, are detailed in Supplementary Table S3. Full details of each counterfactual scenario have been published elsewhere(10, 20, 21). We considered the potential impacts of FOPL implementation solely for changes in sodium intakes. FOPL, front-of-pack labeling; WHO, World Health Organization; AI, Adequate Intake; mg, milligram; CCHS, Canadian Community Health Survey.

#### Meeting sodium reduction targets established by Health Canada (Scenario 3)

Fully meeting Health Canada’s voluntary sodium reduction targets for packaged foods was used as one of the counterfactual scenarios tested in this study. Our research group has previously published methods for this policy modeling approach(10, 37). Briefly, data from a Canadian branded packaged food composition database, the University of Toronto’s Food Label Information and Price database (FLIP 2017), was linked to the Food and Ingredient Details (FID) file in the CCHS-Nutrition 24-hour recall to estimate baseline and counterfactual dietary sodium intakes. The counterfactual scenario was estimated after conducting a systematic ‘reformulation’ of eligible packaged foods to meet Health Canada’s sodium reduction targets^10^ **(Table 2, Supplementary Table S3)**.

#### Implementing ‘high in’ front-of-pack labeling regulations (Scenarios 4-10)

Based on recent FOPL evidence from observational and experimental studies(38-40), we estimated potential changes to dietary sodium intakes(20). These studies estimated changes in the content of nutrients of public health concern (i.e., saturated fats, sodium, and sugars) in food and beverage purchases when a ‘high in’ FOPL was in place. We assumed that changes observed in food and beverage purchases would carry over to dietary intakes, as evidence suggests that documented food purchases can serve as a reasonably precise estimate of overall diet quality(41). We modeled changes in overall food and beverage sodium content based on early evaluations of the Chilean Food Labeling and Marketing Law (-4.7%)(38) (*Scenario 4*), the criteria used by WHO to estimate FOPL cost-effectiveness as part of the *Technical briefing for Appendix 3 of the Global Action Plan for NCDs* (-6.4%)(40) (*Scenario 5*), and a FOPL systematic review and network meta-analysis (-7.8%)(39) (*Scenario 6*), **(Table 2, Supplementary Table S3)**.

Furthermore, we constructed counterfactual scenarios using data from consumer research reporting the proportion of consumers that choose or were willing to choose products with fewer ‘high in’ FOPL symbols(42-47). We first identified foods and beverages in FLIP 2017 similar to those reported in CCHS-Nutrition 2015 that would display at least one fewer ‘high in’ FOPL symbol for nutrients of concern, including sodium. Then, to estimate ‘new’ dietary intakes, we replaced nutritional values of identified foods for a random sample of 30%, 50%, and 70% of *CCHS-Nutrition* adult participants that consume at least one food that would display a ‘high in’ symbol (*Scenarios 7-9*). Additionally, to estimate the potential maximum effect of FOPL, we simulated changes for all adult participants (*Scenario 10*) **(Table 2, Supplementary Table S3)**.

All FOPL counterfactual scenarios tested in this study are based on ‘high in’ FOPL evidence, which is the FOPL system recently approved in Canada. We have previously published methods for all FOPL policy scenarios(20, 21). For this study, we used potential impacts of implementing FOPL only for changes in sodium intakes.

#### Direct healthcare costs

This study followed the perspective of the public healthcare system. The model estimated direct healthcare cost differences between the baseline and the modeled intervention. We used data from the 2010 Economic Burden of Illness in Canada (EBIC)(48), the most recent available and comprehensive national disease-specific costs study. Direct healthcare costs in EBIC 2010 are reported by diagnostic category disaggregated by age/sex groups.

We followed previous published methods for the estimation of direct healthcare costs for each of the modeled diseases(49) (i.e., total and by prevalence case). We prioritized alignment of WHO ICD-10(33) codes between prevalence case data with EBIC 2010 categories, as well as disease definition as closely as possible with the Global Burden of Disease study (2019)(50). Therefore, we first identified the best fitting EBIC diagnostic category by matching ICD-10 codes for each disease under study. We used the EBIC 2010 online tool to produce costs for each EBIC category by sex (male, female) and age group (0-14 years, 15-34, 35-54, 55-64, 65-74, 75+)(51). Direct healthcare costs are comprised of attributable (drug, hospital care – day surgery, hospital care – inpatient, hospital care – other ambulatory care, hospital care – outpatient – clinic, hospital care – outpatient – emergency, and physician care) and unattributable direct costs (other institutions, other professionals, capital, public health, administration, and other health spending).

EBIC diagnostic categories only provide disease-specific estimates for attributable direct costs, but also report total unattributable direct costs; thus, unattributable direct costs were estimated and added to cost estimates per disease. The proportion of each modeled disease’s share to the total attributable direct cost was estimated for each age/sex group using a method developed by Krueger et al(52). Subsequently, this proportion was applied to the total unattributable direct costs to then include these estimates in the total direct healthcare costs for each disease. Costs per disease case were then calculated by dividing estimated direct healthcare costs for each age/sex group by prevalent cases in 2010, yielding the cost per case per year. A decision was made on using prevalent cases for each disease based on their clinical pattern of exerting costs; hence, these were treated as prevalent cases. Lastly, healthcare costs were inflated to 2019 Canadian Dollars (CAD), and inflation factor was estimated using the ‘health care’ sub-index from the Statistics Canada Consumer Price Index(53). Methods, sources, and direct healthcare costs estimations, by each modeled disease, are described in **Table 1** and **Supplementary Tables S4 and S5**.

### 2.3 Sensitivity analysis

Sensitivity analysis was conducted to examine the impact of uncertainty in the discount rate by applying 0% and 3% discount rate, as recommended by the CADTH(22). Additionally, we also used time horizons of 10 and 50 years.

### 2.4 Statistical analysis

The National Cancer Institute (NCI) method(54) was used to estimate Canadian adults’ usual sodium intakes and distributions for all adults and by DRI age/sex group (baseline and counterfactual scenarios). The 1-part (amount only) model was used as zero consumption of sodium was <5%, as recommended by Davis et al(55), which also allowed us to stratify analysis by DRI age/sex groups and to remove outliers for implausible sodium intakes. Confidence intervals and standard error were estimated using the bootstrap balanced repeated replication method (500 replicates). Additionally, we applied sample survey weights (Statistics Canada) to all analyses to ensure nationally representative estimates(35). Statistical analyses were conducted using SAS version 9.4.

After inputting all the required data in PRIMEtime, the model estimated the avoidable ischemic heart disease and stroke incidence cases, healthcare costs savings, and QALYs for each counterfactual scenario, by sex and each disease of interest. Uncertainty intervals (95% UI) were produced using Monte Carlo simulation at 5000 iterations to allow the epidemiological parameters in PRIMEtime to fluctuate randomly following the distributions considered in the model(24).

## 3. Results

### Meeting sodium intake recommendations

Fully meeting Health Canada’s sodium reduction targets was estimated to reduce mean sodium intakes to the national sodium intake recommendations (∼2,300 mg/day; 17% reduction from current levels). This reduction in sodium intake was estimated to prevent 219,490 (95% UI, 73,409 – 408,630) incident cases of IHD, and 164,435 (95% UI, 56,121 – 305,770) strokes. Approximately, 63% of prevented IHD cases were estimated in males (137,188 [95% UI, 44,185 – 259,658]) and 37% in females (82,303 [95% UI, 29,224 – 148,972]), and 57% of prevented strokes were estimated in males (94,522 [95% UI, 31,032 – 179,060]) and 43% in females (69,913 [95% UI, 25,088 – 126,710]). These health gains result in an overall gain of 276,185 QALYs (95% UI, 85,414 – 552,616), with 62% attributed to males and 38% to females. This translates into CAD$ 4,212 million (95% UI, CAD$ 1,303 – CAD$ 8,206) in healthcare cost savings (69% in males, 31% in females) over the lifetime of the 2019 Canadian cohort **(Tables 3 and 4**, **Figure 1 and 2)**.

**Figure 1.**
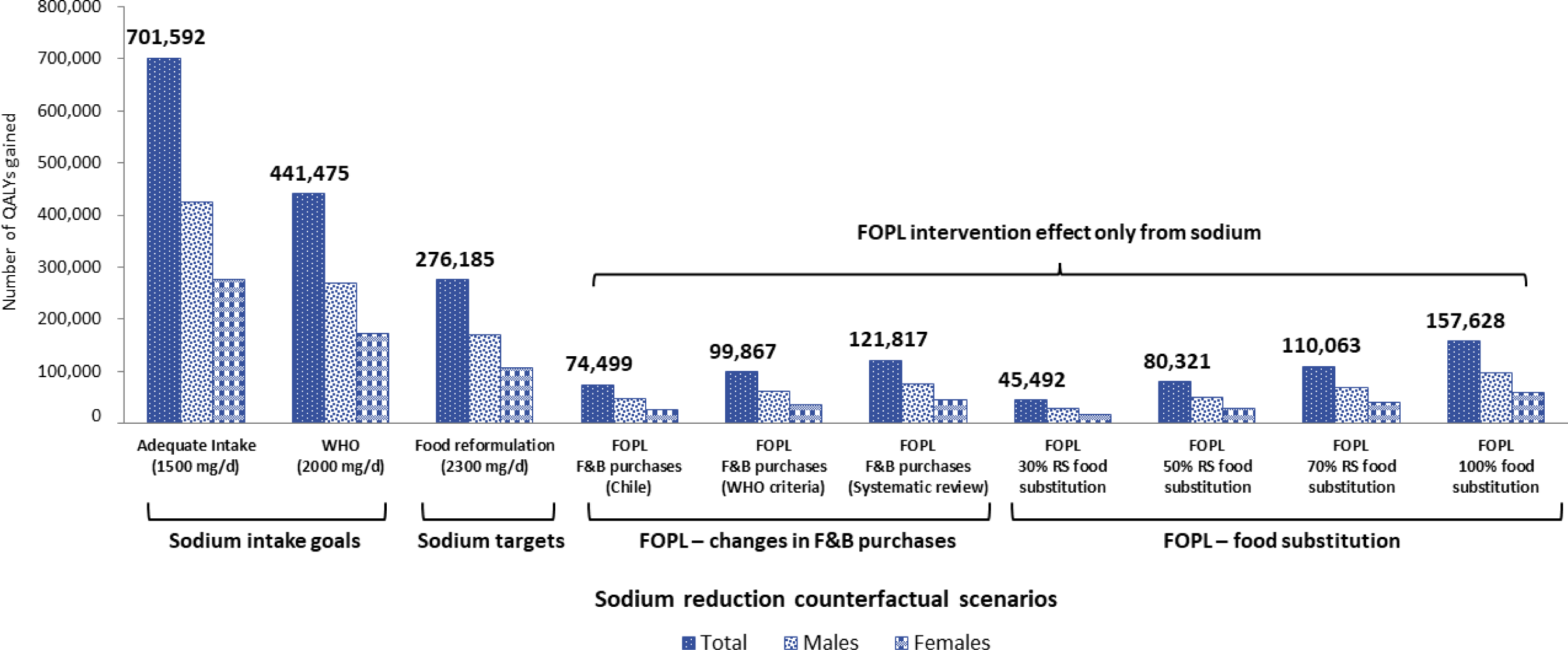
Estimated QALY gained over the lifetime of the 2019 cohort, by sex and sodium reduction strategy. The potential QALYs gained were estimated using the PRIMEtime model(24), with inputs described in Table 1. Full details of each counterfactual scenario have been published elsewhere(10, 20, 21). Sodium intake goal scenarios were based on WHO sodium intake recommendations of 2,000 mg/day and the AI recommendations of 1,500 mg/day. The sodium target scenario was based on fully meeting Health Canada’s voluntary sodium reduction targets for packaged foods. The FOPL–changes in F&B purchase scenarios were based on recent FOPL evidence from observational and experimental studies(38-40). The FOPL– food substitution scenarios were based on consumer research reporting the proportion of consumers that choose or were willing to choose products with fewer ‘high in’ FOPL symbols(42-47). We considered the potential impacts of FOPL implementation solely for changes in sodium intakes. FOPL, front-of-pack labeling; WHO, World Health Organization; AI, Adequate Intake; QALY, Quality-Adjusted Life Year; F&B, food and beverages.

**Figure 2.**
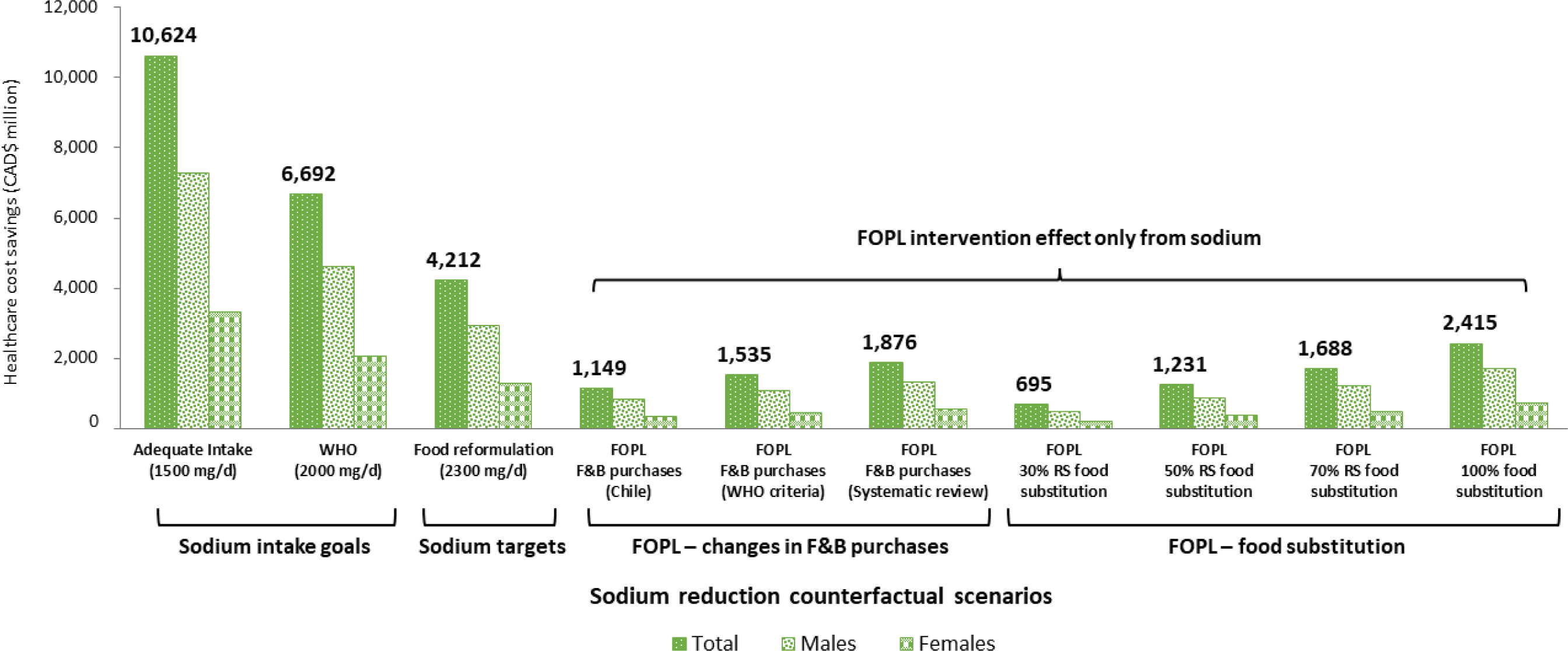
Estimated healthcare cost savings (CAD$ million) over the lifetime of the 2019 cohort, by sex and sodium reduction strategy. The potential healthcare cost savings were estimated using the PRIMEtime model(24), with inputs described in Table 1. Full details of each counterfactual scenario have been published elsewhere(10, 20, 21). Sodium intake goal scenarios were based on the AI recommendations of 1,500 mg/day and the WHO sodium intake recommendations of 2,000 mg/day. The sodium target scenario was based on fully meeting Health Canada’s voluntary sodium reduction targets for packaged foods. The FOPL–changes in F&B purchase scenarios were based on recent FOPL evidence from observational and experimental studies(38-40). The FOPL–food substitution scenarios were based on consumer research reporting the proportion of consumers that choose or were willing to choose products with fewer ‘high in’ FOPL symbols(42-47). We considered the potential impacts of FOPL implementation solely for changes in sodium intakes. FOPL, front-of-pack labeling; WHO, World Health Organization; AI, Adequate Intake; QALY, Quality-Adjusted Life Year; F&B, food and beverages.

**Table 3.**
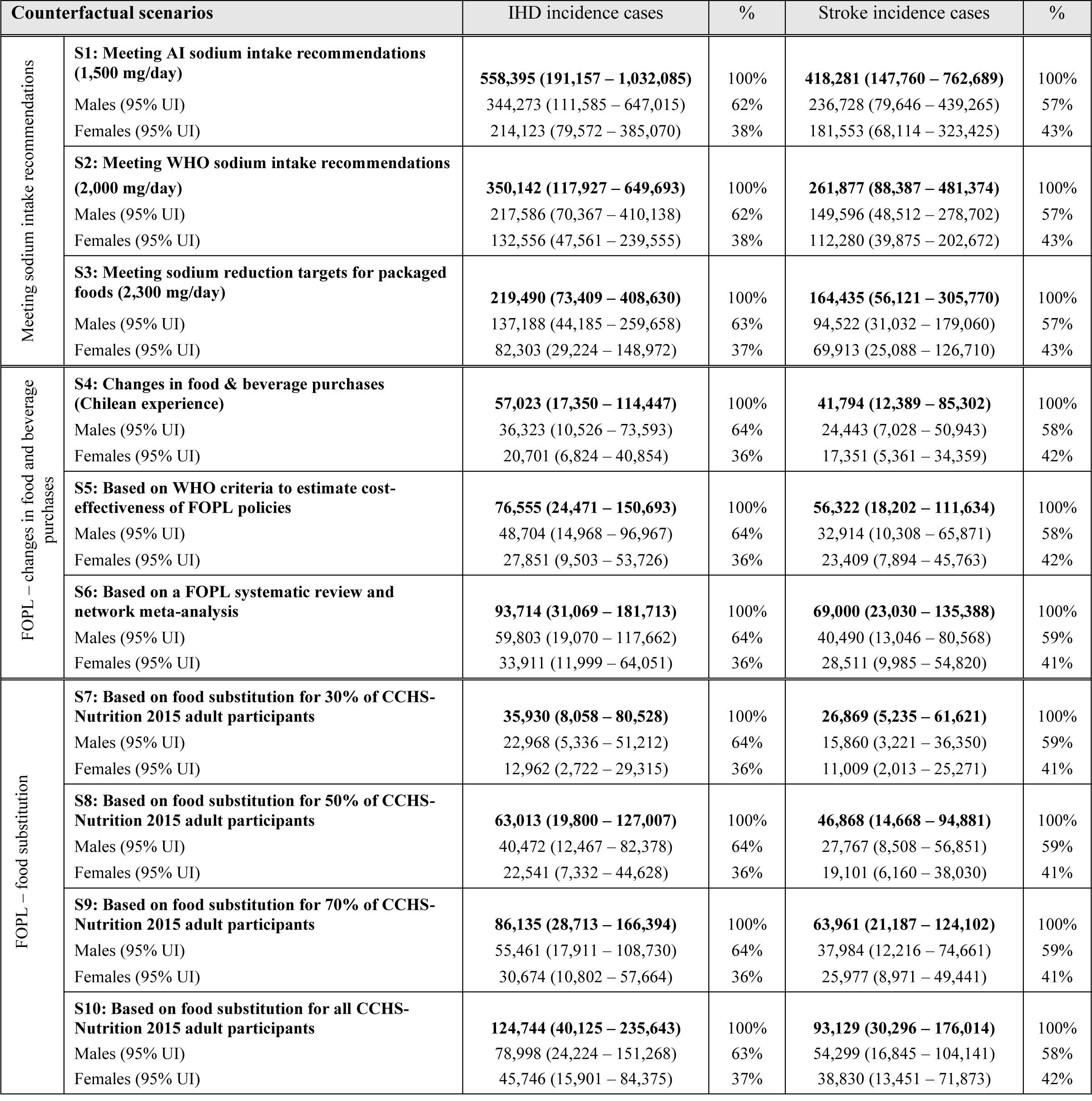
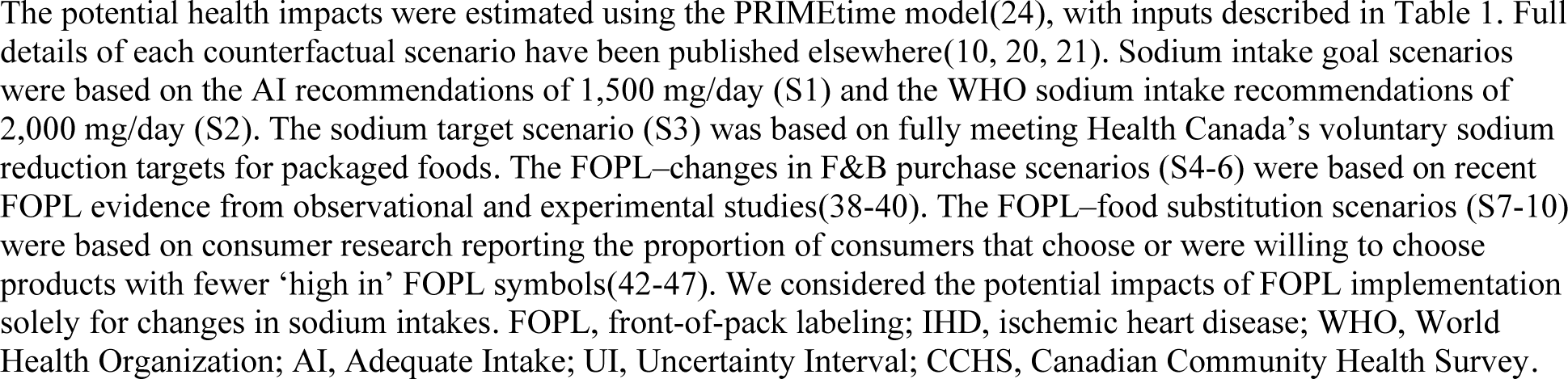
Prevented disease incident cases over the lifetime of the cohort, by sex and sodium reduction strategy.

**Table 4.**
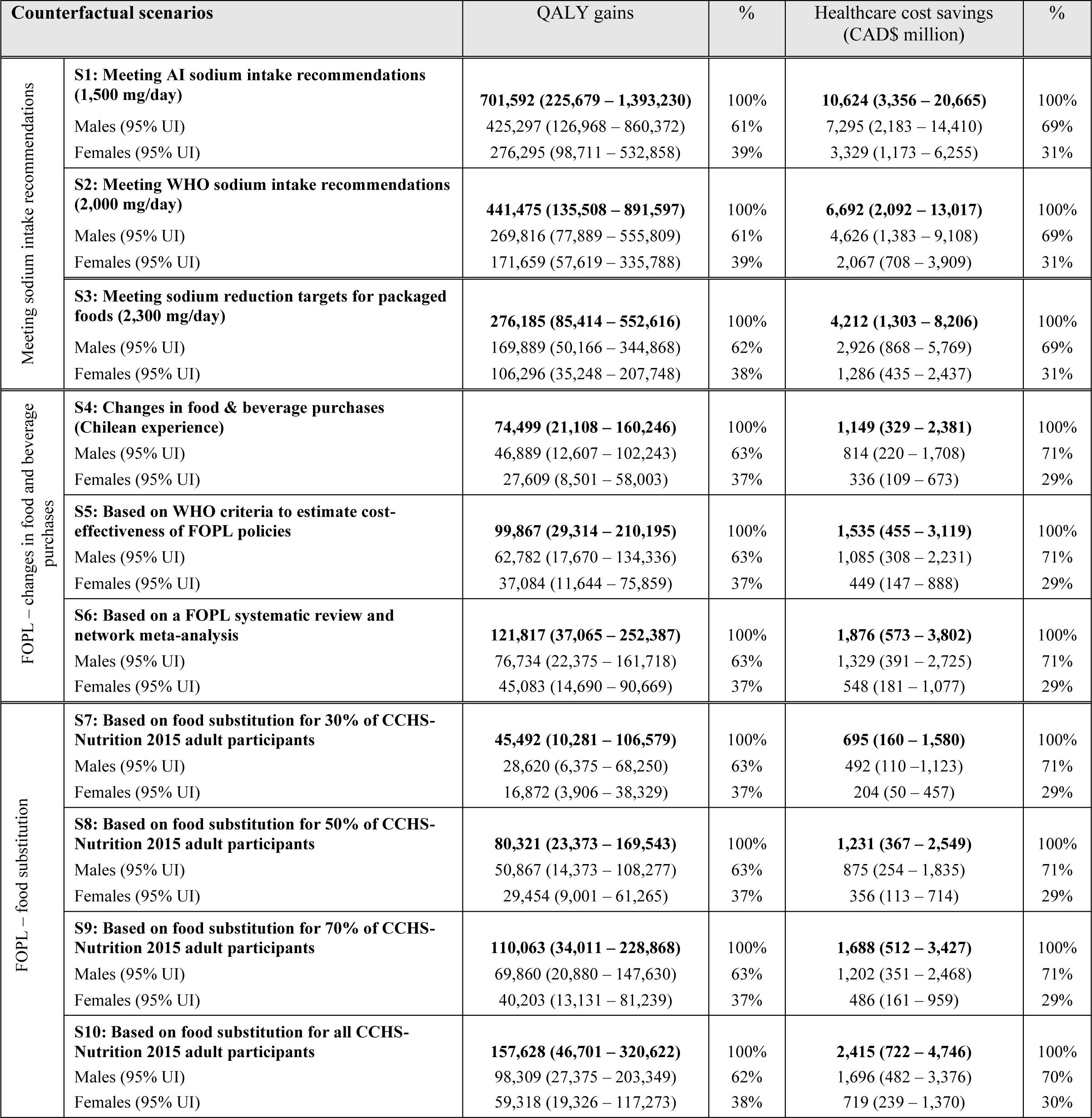
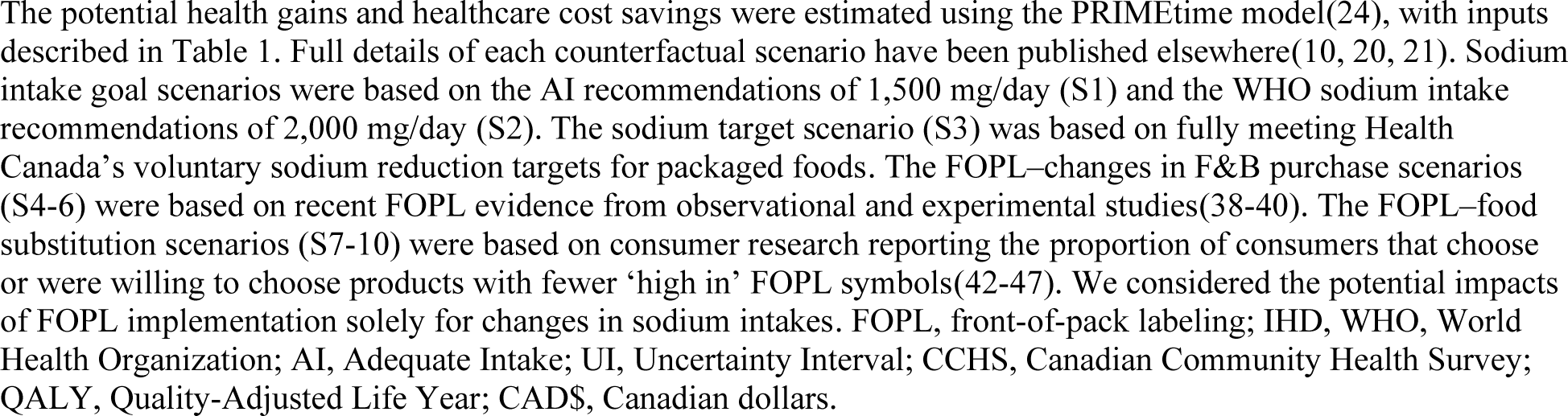
Estimated health gains and healthcare cost savings over the lifetime of the cohort, by sex and sodium reduction strategy.

Greater health and healthcare cost gains were estimated if Canadians were to meet WHO’s population-level sodium intake recommendations (2,000 mg/day) and the Adequate Intake recommendation (1,500 mg/day) **(Tables 3 and 4**, **Figure 1 and 2)**.

### Implementing ‘high in’ front-of-pack labeling regulations

Sodium intake reduction through the implementation of FOPL regulations, as suggested by early evaluations of the Chilean FOPL regulations, has the potential to prevent 57,023 (95% UI, 17,350 – 114,447) incident cases of IHD, and 41,794 (95% UI, 12,389 – 85,302) strokes.

Approximately 64% of prevented IHD cases were estimated in males (36,323 [95% UI, 10,526 – 73,593]) and 36% in females (20,701 [95% UI, 6,824 – 40,854]), and 58% of prevented strokes were estimated in males (24,443 [95% UI, 7,028 – 50,943]) and 42% in females (17,351 [95% UI, 5,361 – 34,359]). These health gains result in an overall gain of 74,499 QALYs (95% UI, 21,108 – 160,246), with 63% attributed to males and 37% to females. This translates into CAD$ 1,149 million (95% UI, CAD$ 329 – CAD$ 2,381) in healthcare cost savings (71% in males, 29% in females) over the lifetime of the 2019 Canadian cohort **(Tables 3 and 4**, **Figure 1 and 2)**.

Greater health and healthcare cost gains were projected based on the criteria used by WHO to estimate FOPL cost-effectiveness and results from a FOPL systematic review and network meta-analysis. An increase of 34% and 64% in all outcomes was estimated, respectively, versus the results based on evaluations of the Chilean FOPL **(Tables 3 and 4**, **Figure 1 and 2)**.

Sodium intake reduction through consumer food substitution – specifically, when consumers choose food products with fewer ‘high in’ FOPL symbols – has the potential to prevent between 35,930 (95% UI, 8,085 – 80,528) and 124,744 (95% UI, 40,125 – 235,643) cases of IHD, and between 26,869 (95% UI, 5,235 – 61,621) and 93,129 (95% UI, 30,296 – 176,014) strokes. This results in QALY gains ranging from 45,492 (95% UI, 10,281 – 106,579) to 157,628 (95% UI, 46,701 – 320,622), and healthcare cost savings ranging from CAD$ 695 million (95% UI, CAD$ 160 – CAD$ 1,580) to CAD$ 2,415 million (95% UI, CAD$ 722 – CAD$ 4,746) over the lifetime of the 2019 Canadian cohort **(Tables 3 and 4**, **Figure 1 and 2)**.

All sodium reduction strategies tested in this study were cost saving from the public healthcare system perspective. Estimated benefits were greater for males than females in all scenarios tested.

### Sensitivity analysis

We examined the impact of uncertainty in the discount rate by applying discount rates of 3% and 0%. When a 3% discount rate was applied to both health and healthcare cost outcomes, a decrease of 44% and 39% were observed on average for QALYs and healthcare costs, respectively, in all scenarios. Conversely, when a 0% discount rate was applied, an increase of 96% and 78% were observed on average for QALYs and healthcare costs, respectively, in all scenarios (See **Supplementary Table S10**).

Furthermore, using time horizons of 10 and 50 years, the model predicted that 7% and 76%, respectively, of estimated lifetime healthcare cost savings would occur within those time horizons. For QALYs gained, the model estimated that 4% and 67%, respectively, of estimated lifetime gains would occur within those same time horizons (See **Supplementary Table S11**).

## 4. Discussion

This study estimated the potential impact of reducing sodium intake on the incidence of ischemic heart disease and stroke cases, savings in healthcare costs, and QALYs gains. Our results demonstrate that reducing sodium intakes through population-level strategies, such as implementing sodium reduction targets and ‘high in’ FOPL regulations, has the potential to improve health outcomes and reduce healthcare expenditures in Canada. This adds to current evidence, generated by simulation modeling studies, showing that sodium reduction strategies can potentially improve health outcomes and save costs to the health system, globally(29, 56-62) and in Canada(10, 19).

To the best of our knowledge, this is the first study estimating the potential health (QALYs) and healthcare costs of current sodium reduction initiatives in Canada. We tested ten counterfactual scenarios aiming to reduce population sodium intake. These included achieving recommended sodium intakes levels (2,000 and 1,500 mg/day); meeting sodium reduction targets for packaged foods established by Health Canada (food reformulation, 2,300 mg/day); and estimated changes in sodium intake resulting from implementing the recently approved ‘high in’ FOPL regulations. As expected, our results showed the greatest health gains and healthcare cost savings from meeting population-level sodium intake recommendations of 2,000 mg/day (441,475 QALYs; CAD$ 6,692 million in healthcare costs savings) and 1,500 mg/day (701,592 QALYs; CAD$ 10,624 million in healthcare costs savings). Our study sheds light on how far Canadians are from achieving these maximum benefits with current sodium reduction policy initiatives.

For instance, for the policy initiatives tested in this study (i.e., meeting sodium reduction targets and implementing ‘high in’ FOPL regulations), most health and healthcare costs gains were attributed to fully meeting Health Canada’s sodium reduction targets (276,185 QALYs; CAD$ 4,212 million in healthcare costs savings). Greater benefits were observed for males than females, in all scenarios tested, which could be attributed to differences in sodium consumption and CVD burden among Canadians males and females(9, 10, 63). Unfortunately, progress evaluations have shown that compliance with the voluntary sodium reduction targets has been limited, with only 14% of food categories meeting the targets by 2017(14), indicating that to meet targets by 2025(13) faster progress is needed. A robust government-led monitoring system and higher compliance from the food industry could contribute towards increasing sodium reduction progress in Canada. The consequences of not meeting established sodium reduction targets for processed foods represents an important missed opportunity to generate substantial health and healthcare costs gains in Canada.

Canada could benefit from other countries experiences that have implemented mandatory sodium reduction targets. For instance, South Africa’s legislation setting mandatory maximum sodium levels in foods has led to a decrease of 1.15 g/day in salt intake (∼460 mg/day of sodium) in a four-year period (2015-2019). Interestingly, similar sodium reduction levels were estimated for our scenario of fully meeting sodium reduction targets(64), which indicates the feasibility of this strategy. This also highlights the importance of changing the voluntary nature of sodium reduction targets in Canada to mandatory sodium reduction targets. More recently, the WHO released the *WHO global sodium benchmarks for different food categories*(65). This set of sodium reduction targets includes 18 main food categories and 97 food subcategories. Greater benefits would be expected from meeting these targets in Canada, given that for several common food categories WHO targets are more stringent than Health Canada’s sodium reduction targets. In Australia, it has been estimated that meeting WHO benchmarks, as opposed to meeting Australian sodium reduction targets, could prevent or delay nearly three and a half times as many diet related NCD deaths(62).

The potential impact of FOPL regulations was estimated through two pathways. The first was based on recent FOPL evidence from observational and experimental studies(38-40) that estimated changes in the content of nutrients of public health concern, including sodium, in food and beverage purchases when a ‘high in’ FOPL was in place. Among these scenarios is one based on early evaluations of the Chilean FOPL (S4), which most likely captured consumer behavior change and initial industry-driven food reformulation. The health and healthcare cost gains estimated from this scenario represent approximately 27% of the gains estimated from meeting Health Canada’s sodium reduction targets. The second pathway was based on consumer research reporting the proportion of consumers that choose or were willing to choose products with fewer ‘high in’ FOPL symbols(42-47), which would only capture consumer behavior change. The estimated benefits from these scenarios range from 16% to 57% of the benefits estimated from meeting Health Canada’s sodium reduction targets.

The differences in FOPL scenarios tested are reflected in our estimates. We observed greater benefits from changes observed in food and beverage purchases in the presence of FOPL (S4-6) than from food substitution as a response of FOPL (S7-9). It is worth noting that our study focused solely on benefits from reducing sodium intake as a result of implementing FOPL in Canada. Therefore, it is expected that implementation of FOPL will result in greater health and healthcare cost benefits, as it also targets other nutrients-of-concern (i.e., sugars and saturated fats) not accounted for in our estimates. For instance, a recent study from our research group estimated that 15% of diet related NCD deaths that could be averted or delayed due to implementation of FOPL in Canada were attributed to sodium(21).

A previous Canadian modeling study estimated potential heath and healthcare cost benefits, over a 50-year time horizon, from reducing sodium intake levels by 1,500 mg/day(19). They estimated benefits of 1,021,458 QALYs and CAD$16,805 million in healthcare cost savings(19). In comparison, the largest reduction we tested was a reduction in sodium intake of 1,258 mg/day to achieve the AI recommendations (1,500 mg/day). Under this scenario we projected benefits of 471,748 QALYs and CAD$ 8,083 million in healthcare cost savings over a 50-year time horizon. Differences likely stem from variations in the magnitude of the tested sodium reduction intake (reductions of 1500 mg/day vs. 1,258 mg/day), differences in methodology used to estimate healthcare costs, improved management of SBP in Canada, and slight reductions observed in the burden of CVDs in Canada. In the US a sodium reduction of 1,200 mg/day has been predicted to gain between 194,000 and 392,000 QALYs and save between $10,000 to $24,000 million in healthcare costs(66).

This study has limitations and strengths that need to be considered in the interpretation of our results. First, scenario modeling is an analytical technique; hence, results are as appropriate and generalizable as the data, assumptions and constraints that are applied to the mathematical model(67). To mitigate this, we used Canadian–specific and nationally representative data as inputs for the model. Additionally, our counterfactual sodium reduction intervention scenarios are based on Canadian sodium reduction targets and the most recent available evidence on the impact of FOPL regulations. We followed the perspective of the public healthcare system; hence, the costs of implementing the sodium reduction interventions were not considered. Although population-wide sodium reduction interventions in Canada were estimated to cost CAD$2.02 per person annually (Canadian population in 2019: 38 million approximately), these costs would be minimal in comparison with savings in healthcare costs(19, 68). These costs included government led industry agreements to reduce sodium in packaged foods, government monitoring of industry compliance, and public health campaigns(19, 68). We also did not consider societal gains in our modeling, such as productivity gains from preventing premature deaths or disease burden.

Furthermore, we focused on the potential health effects of reducing sodium intake, specifically health effects from IHD and stroke, the leading causes of death globally. However, recent studies have estimated potential effects of reducing sodium on other diseases such as chronic kidney disease and stomach cancer(58, 59, 69, 70). Therefore, our estimations of the potential benefits of reducing population-level sodium intake are conservative, considering that reducing sodium intake could also have positive effects on other diseases. Lastly, this study used data for the entire Canadian population without considering equity aspects. Future research can expand our work by examining the differential impacts of reducing sodium intake among diverse subgroups.

This study also has strengths to consider. Dietary sodium intake data was estimated using data from a nationally representative sample of the Canadian population (CCHS-Nutrition 2015). Surveys of this kind are usually prone to biases related to misreporting due to recall bias. Nevertheless, CCHS-Nutrition 2015 used the Automated Multiple Pass Method to reduce the impact of such reporting errors. We used the NCI method to estimate usual sodium intakes and adjusted for age, sex, dietary misreporting status, weekend/weekday, and sequence of dietary recall. For modeling we used the PRIMEtime model, a robust, validated and widely used proportional multi-state lifetable model(23, 24). Strengths and limitations of the model have been previously published(24). We also conducted sensitivity analysis to examine the impact of uncertainty in the discount rate by applying 0% and 3% discount rates, as has been recommended(22). As expected, variations in discount rates, both higher and lower than our main analysis (1.5% discount rate), had a significant impact on health and healthcare cost outcomes.

Our findings provide evidence for policymakers and stakeholders of the potential benefits of reducing sodium intake through strategies widely discussed in Canada that target packaged foods. This is especially relevant because packaged foods are the main source of sodium intake for Canadians(9). Multicomponent strategies have proven to be more effective and are necessary for reaching the national and global sodium recommendations(11, 71). Therefore, in addition to moving from voluntary to mandatory sodium reduction targets for packaged foods and implementing ‘high in’ FOPL regulations, other initiatives should be considered for Canadians to meet population-level sodium intake recommendations (2,000 and 1,500 mg/day). These could include setting sodium reduction targets for restaurant foods, restricting marketing of foods ‘high in’ sodium, enabling healthy food environments in public settings by providing lower sodium options, regulating the digital food environment to require nutritional information, encouraging the use of low-sodium, potassium-enriched salt, and adopting approaches such as social marketing to develop behavior change communication and mass media campaigns(11, 72, 73). A combination of strategies, as has been recommended by the WHO’s *SHAKE Technical Package for Salt Reduction*(11), would be of most benefit from a public health standpoint.

## 5. Conclusions

Reducing population-level sodium intakes is feasible and has the potential to improve health outcomes and save healthcare costs in Canada. From the interventions tested most health and healthcare costs gains were attributed to fully meeting sodium reduction targets, which highlights the importance of changing the voluntary nature of these targets to mandatory. A combination of strategies, mandatory sodium reduction targets and implementation of the ‘high in’ FOPL symbol would be of most benefit from a public health standpoint. Thus, policymakers and stakeholders need to strengthen, and coordinate actions aimed at reducing sodium levels in the Canadian food supply. This is crucial for improving health outcomes and saving healthcare costs. Future studies could leverage this work to evaluate potential variations in effects among diverse subgroups in Canada.

## Authors’ contributions

NF, AJ, and MRL conceptualized the study design; NF, AJ, BAC, LC, EN and MRL interpreted the findings; NF conducted the study, wrote the original draft, and performed the statistical analysis. All authors critically reviewed and approved the final manuscript.

## Acknowledgments

The authors would like to thank Professor Peter Scarborough, University of Oxford, for allowing us to use the PRIMEtime model and discussing its application; as well as current and past L’Abbe lab members who worked on various aspects of the FLIP database, especially Anthea Christoforou who worked and published data on the sodium reformulation scenario used in this manuscript. Furthermore, the authors would like to acknowledge the assistance of Arantxa Bonifaz Rosas with the collection of Canadian healthcare cost data.

## Data Availability Statement

Analytic code (R) can be made available to researchers upon request to the author. *Canadian Community Health Survey-Nutrition 2015 Public Use Microdata File (PUMF)* data is publicly and freely available without restriction at Statistics Canada, https://www150.statcan.gc.ca/n1/en/catalogue/82M0024X

Canadian population demographics, healthcare costs and epidemiology data associated with ischemic heart disease and stroke were obtained from publicly available sources – also detailed in the main manuscript.

https://www150.statcan.gc.ca/t1/tbl1/en/tv.action?pid=1710000501

https://www150.statcan.gc.ca/t1/tbl1/en/tv.action?pid=1310070901

https://www150.statcan.gc.ca/t1/tbl1/en/tv.action?pid=1310014701

https://health-infobase.canada.ca/ccdss/data-tool/

https://cost-illness.canada.ca/custom-personnalise/national.php

